# Cost-effectiveness of Targeted Next Generation Sequencing for TB drug-resistance testing as an alternative to the standard of care in South Africa

**DOI:** 10.1101/2025.03.13.25323893

**Authors:** Alexandra de Nooy, Shaheed Vally Omar, Tom Ockhuisen, Alice Zwerling, Suvesh Shrestha, Anita Suresh, Shaukat Khan, Rebecca E. Colman, Swapna Uplekar, Timothy C. Rodwell, Nazir Ismail, Kyra Grantz, Sarah Girdwood, Brooke E. Nichols

## Abstract

**Background:** South Africa faces emerging resistance to key TB drugs, including bedaquiline. Phenotypic drug susceptibility testing (pDST), the current reference standard for bedaquiline DST, while accurate has long turnaround times. Targeted next-generation sequencing (tNGS) offers a comprehensive alternative to pDST, potentially delivering faster results. However, its advantages must be weighed against differences in implementation cost and test accuracy.

**Methods:** We used a decision tree model to evaluate the cost-effectiveness of tNGS against the standard of care (SOC) in South Africa at different levels of tNGS decentralization (1, 3, 4, or 6 sites). Key outcomes considered were survival rates, time to a correct resistance profile, duration of infectiousness, and disability-adjusted life years (DALYs). Sensitivity analyses assessed the impact of drug resistance prevalence, tNGS sensitivity, and improved DST access on DALYs and incremental cost per DALY averted.

**Results:** tNGS averted 408 DALYs and returned a greater number of correct resistance profiles (90.7%) as compared to the SOC (87.7%). Based on model and scenario assumptions for South Africa, tNGS returned results with a reduced turnaround time and averted 96 years of infectious time. Centralized tNGS was determined to be cost-saving relative to the SOC, however decentralization of tNGS resulted in higher incremental costs per DALY averted ($671-$2,454). tNGS performance relative to the SOC improved at higher bedaquiline resistance prevalence and when tNGS sensitivity increased. Access gains through tNGS increased the number of DALYs averted and decreased the respective incremental cost per DALY averted for decentralized scenarios.

**Conclusions:** Centralized tNGS testing is likely to be cost-saving in South Africa and decentralised tNGS would result in higher costs but could be cost-effective under current assumptions. Additionally, tNGS has the potential to reduce DALYs, shorten result turnaround times, and decrease infectious duration while improving the percentage of individuals receiving correct DST results.

## INTRODUCTION

Tuberculosis (TB) remains one of the deadliest infectious diseases globally, with an estimated 10.8 million cases and 1.25 million deaths reported in 2023^1^. South Africa faces a particularly heavy burden of TB, being included in all three of the World Health Organization’s (WHO) high-priority lists: 30 high-TB burden countries, 30 countries with high TB/HIV coinfection, and 30 countries with high multidrug resistant (MDR) and/or rifampicin resistant (RR-) TB ^1^. In 2023, South Africa reported approximately 193 000 notified cases of TB out of an estimated 270 000, with 5.1% of these cases being RR-TB ^2–4^. Beyond RR-TB, resistance to other critical drugs, such as isoniazid, fluroquinolones, linezolid, and bedaquiline, also poses significant challenges ^2,5^. Drug-resistant TB (DR-TB) is associated with worse outcomes-including higher mortality and increased loss-to-follow-up rates-compared to drug-sensitive TB (DS-TB) ^6^. These worse outcomes often stem from barriers to timely diagnosis and treatment, including access to accurate and rapid drug resistance testing, and the historically longer, more toxic treatment regimens for DR-TB compared to DS-TB ^7–9^.

In South Africa, the current standard of care (SOC) for drug susceptibility testing (DST) is initiated following the detection of RR-TB returned alongside the initial TB diagnosis^10^. Following a positive RR-TB result, it is recommended that a sample is sent for additional drug-resistance testing using the Xpert MTB/XDR^11^ (to assess resistance to drugs such as isoniazid and fluroquinolones) and phenotypic drug susceptibility testing (pDST) for bedaquiline and linezolid ^12^. Any additional drug resistance identified through these tests necessitates modifications to the initial RR/MDR TB regimen, which is comprised of bedaquiline, pretomanid, linezolid and levofloxacin (BPaL-L).

Currently, resistance to three key drugs in the BPaL-L regimen-bedaquiline, linezolid and pretomanid -can only be determined through pDST ^12^. However, pDST in South Africa faces significant challenges, including long turnaround times (TAT) (median time of 56 days) and limited access (with only ∼43% of rifampicin resistant individuals receiving bedaquiline pDST results) ^2,4^. Consequently, many individuals with additional drug resistance may remain on incorrect regimens for extended periods, either until pDST results are returned or treatment failure is identified. This delay contributes to worse patient outcomes and increases the risk of onward transmission of MDR-TB^13^. These challenges underscore the urgent need for additional tools to support or enhance DST.

Targeted next-generation sequencing (tNGS) is one such tool under consideration for implementation and offers several potential advantages over existing DST methods ^14^. Unlike other tools, tNGS can detect resistance to almost all drugs used in TB treatment in a single test directly from the collected specimen ^14^. Furthermore, it allows for the rapid inclusion of new resistance markers as they are identified, offering adaptability that many current diagnostic methods lack ^15^. Although the TAT for tNGS could be shorter than that of pDST, it depends on operational factors such as batch size, sample volume, run frequency, and technical expertise ^16^. As such, being able to quickly and easily conduct DST through tNGS would be particularly important for drugs like bedaquiline and linezolid, which are key components of the BPaL-L regimen.

Recently, work by Shrestha et al ^17^ indicated that tNGS could be cost-effective across different contexts, particularly where there is no comprehensive DST already in place. Specifically, in South Africa, tNGS was found to be cost-effective compared to the SOC at a willingness-to-pay threshold of three times the GDP per capita and using a sensitivity to bedaquiline for tNGS of 68%. Since the time of this analysis, additional studies have further evaluated the sensitivity of tNGS for detecting resistance to various drugs. Colman et al reported tNGS sensitivity values for bedaquiline at 83.9% and 46.2% for linezolid ^18^, both of which are lower than the respective sensitivity values reported for pDST ^15^, but, the sensitivity for bedaquiline was notably higher than the 68% sensitivity initially reported by the World Health Organization (WHO) ^19^. Thus, the potential TAT and test completion benefits of tNGS must be weighed against its lower sensitivity. Additionally, the cost implications of integrating tNGS into the DST algorithm and national testing programme require careful consideration.

In this analysis we used a decision tree model, adapted from Shrestha et al ^17^, to compare tNGS implementation scenarios (at different levels of decentralisation) against the SOC. The analysis evaluated cost-effectiveness, the accuracy of resistance profiling, TAT, and the potential effect on reducing overall time in which transmission could occur.

## METHODS

### Developed model and relevant outcomes

The original model by Shrestha et al ^17^ was built in TreeAge Pro Healthcare ^20^ and considered the cost-effectiveness of tNGS against the SOC (Xpert XDR and pDST). The adapted model for this work was developed in R Studio (RStudio 2022.07.2 ^21^) and follows the form of a decision tree which maps out, for each individual with a specific resistance profile, the testing process per algorithm, result return, treatment regimen and finally the likelihood of treatment initiation, completion and survival. The model simulates each DST algorithm for a population of 10,000 RR-TB individuals (an approximate value for the number of RR-TB cases historically seen in South Africa ^22^), with additional drug resistance profiles randomly allocated to individuals based on prevalence estimates. Many of the values used to parameterise the decision tree follow those in Shrestha et al ^17^, but several have been updated based on newer data and additional discussion with in-country experts. A full list of parameter values and corresponding sources is provided in Appendix: Table S1.

To evaluate each DST algorithm, several outcomes are compared. The first outcome is survival rate. In this model, assumptions on mortality broadly follow those in Shrestha et al ^17^, however, an additional assumption has been made with respect to individuals correctly switching to individualised regimens (Appendix: Table S2). Here we now account for additional mortality that might occur if the return of results (and subsequent changing of regimen) is delayed, resulting in patients spending a longer time on ineffective treatment. Additionally, the percentage of resistance profiles correctly determined and returned is calculated. Two time-related outcomes are also estimated: the time to a correct resistance profile being returned, and the estimated infectious time. The time to a correct resistance profile accounts for the expected TAT per algorithm. This is estimated to be 10.5 days for tNGS based on a 7-day in-laboratory sequencing TAT and an additional 3.5 days for sample transport and result review. This time meets the desired requirements of the National TB Reference Laboratory NICD/NHLS (NICD) to keep the total TAT within 14 days. For the SOC, TAT is estimated to be 56 days based on the median reported time for bedaquiline pDST results being returned^2^. Finally, time penalties of 84 days (based on 3-month culture check for treatment failure^23^) are included for missing or incorrect resistance results. Infectious time, calculated both in total and for each resistance profile, represents the estimated time a patient remains infectious between diagnosis and the initiation of effective treatment. The parameters and equations used to estimate this duration for each relevant group, categorized by resistance profile and treatment outcome, are detailed in Appendix: Table S3.

### Cost-effectiveness analysis

In this analysis we considered different potential implementation rollout strategies of tNGS in South Africa in terms of the number of sites conducting testing. In this we compared one centralised site against three decentralised scenarios with 3,4 and 6 sites respectively as requested by the NICD. For each implementation scenario the total cost was calculated and, in conjunction with the relevant DALYs, was used to calculate the incremental cost per DALY averted between tNGS scenarios and the SOC. DALYs were calculated based on years of life lost (YLL) for each modelled death and years of life lived with a disability (YLD) ^24^. Detailed calculations for DALYs by resistance profile and treatment outcome group are provided in Appendix: Table S4.

The cost analysis estimated costs associated with the SOC and tNGS DST algorithms (Appendix: Table S5-S7). For tNGS, cost estimates were based on projected volumes for RR-TB cases (10,000 individuals), the number of implementation sites (1,3,4, or 6) and the sequencing instrument and method used for tNGS (Illumina’s NextSeq1000^25^ and GenoScreen’s Deeplex Myc-TB test^26^). The degree of decentralization influenced the per-test costs: increased decentralization required more instruments and reduced operational efficiency due to smaller batch sizes at low-volume sites (Appendix: Table S7). In addition, we distinguished between two separate objectives when estimating the cost per test for tNGS: minimizing TAT and minimizing cost. In the minimize TAT scenarios, instruments are run at set intervals (at least one run per week) to maintain a low TAT, despite resulting in higher costs. In the minimize cost scenarios, instrument utilization is maximized by running instruments only when fully loaded with samples, which may involve up to 4 weeks of wait time, depending on site-specific test volumes. However, TAT is critical, with sequencing for tNGS needing to be completed within a maximum of 7 days to align with the NICD’s stipulated 14-day TAT while still allowing time for sample transportation between sites. Where TAT was within 7 days, costs were minimized, and in other scenarios higher costs were accepted to ensure TAT did not exceed 7 days. Given South Africa’s extensive sample transport system, additional transport costs for decentralisation are negligible and it is assumed that greater levels of decentralisation do not affect require longer TAT. For the SOC strategy, costs included consumables, equipment, staff, and energy expenses for both pDST and Xpert XDR testing All costs were adjusted for inflation (where necessary) to 2024 values and converted to U.S. dollars using an exchange rate of R18.04/USD.

### Sensitivity analyses

Given that prevalence of drug resistance changes over time, it is important to evaluate the performance of tNGS relative to the SOC under varying prevalence levels. In this analysis, the focus was on bedaquiline, a core drug of the BPaL-L regimen, which is less frequently tested for resistance compared to drugs like rifampicin and fluroquinolones. Resistance to bedaquiline is also already more prevalent as compared to resistance to linezolid ^2^. In the sensitivity analysis, the prevalence of bedaquiline resistance was varied from baseline by applying a 0.5 to a 4-fold increase to the baseline bedaquiline resistance prevalence parameter values. Additionally, given the variability in sensitivity values reported in the literature for bedaquiline resistance detection (for example lower bounds of 68% have previously been reported by the WHO and 83.9% by Colman et al ^15,18^) the performance of tNGS against the SOC was compared across a range of tNGS bedaquiline sensitivity values (60%-100%). These analyses were combined and the total DALYs determined in each scenario.

Further, while the base model assumes no difference in access to DST between tNGS and SOC, tNGS has the potential to improve DST access and result availability. This improvement could stem from tNGS offering more reliable TAT, enabling clinicians to know when tNGS results are available or prompting additional DST requests if results are delayed or not returned. To assess this, the analysis considered increases in DST coverage attributable to tNGS, ranging from 0%-20%, and calculated the total DALYs averted under each scenario. This was conducted for each level of tNGS decentralisation, and the subsequent incremental cost per DALY averted, for each combination of access gain and decentralisation level), was determined to assess impact on cost-effectiveness.

## RESULTS

tNGS increases the number of individuals receiving DST (∼7.5%) but has marginal gains with respect to initiating and completing treatment and overall survival (∼0.2% increase) relative to the SOC (Figure 1a). Whilst tNGS correctly returns a greater percentage of results to those with rifampicin mono resistance and fluroquinolone resistance as compared to the SOC, for all other resistance profiles the SOC outperforms tNGS (Figure 1b). Overall, the total percentage of results correctly returned is 90.7% and 87% for tNGS and the SOC respectively (Figure 1a).

**Figure 1:**
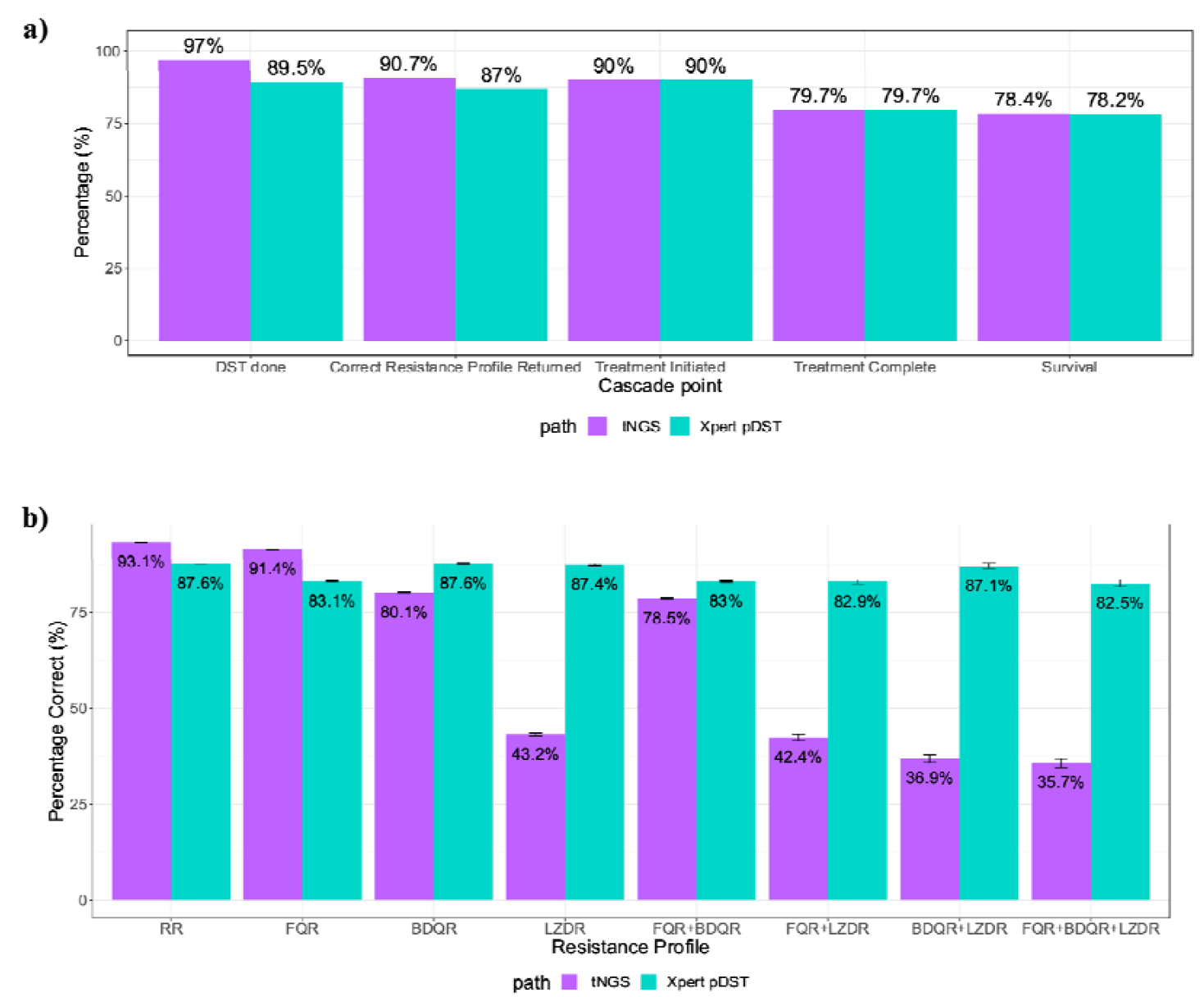
TB cascade and resistance profile comparison for drug-susceptibility testing through tNGS and the standard of care (GeneXpert XDR and pDST). Panel A: TB drug-resistance testing and treatment cascade per testing strategy. Panel B: Comparison of percentage resistance profiles correctly returned per profile and testing strategy

The tNGS algorithm returns results more rapidly and reduces infectious time as compared to the SOC algorithm. The time to a complete drug-resistance profile being returned is estimated, on average, at 59.6 days per individual in the SOC and 17.3 days per person when using tNGS. Consequently, the tNGS algorithm is estimated to reduce infectious time during DST with an estimated 5,388 years of infectious time compared to 5,485 years for the SOC-a reduction of 97 years of infectious time.

With respect to costs, the per-test cost of GeneXpert XDR was estimated at $39.90 ^27^, while pDST was $97.00, resulting in a combined per-test cost of $137 under the SOC algorithm (Appendix: Table S5). For the tNGS algorithm, per-test cost varied based on the degree of decentralization: $102 per test for 1 site, $148 for 3 sites, $171 for 4 sites, and $215 for 6 sites (Table 1, Appendix: Table S6). Overall, the SOC resulted in a higher number of DALYs (70,893) compared to the tNGS testing strategy (70,485, resulting in 408 DALYs averted with tNGS (Table 1). When tNGS testing is conducted at a single site it is cost-saving relative to the SOC, however as tNGS testing is decentralized to more sites, the incremental cost per DALY averted rises (ranging from $671 to $2453 per DALY averted).

**Table 1:** Results of cost-effectiveness analysis.

| **DST Algorithm**<br>**(cost per test)** | **Total Cost (\$)**<br>**(Range)** | **Total DALYs**<br>**(Range)** | ***Incremental cost per***<br>***DALY averted***<br>**(Range)** |
| --- | --- | --- | --- |
| Xpert + pDST<br>(\$137) | \$1.3m<br>(\$1,333,883 - \$ 1,334,147) | 70,893<br>(70,840 – 71,077) | - |
| tNGS<br>(1 site, \$102) | \$1.1m<br>(\$1,107,951 – \$1,108,319) | 70,485<br>(70,431 – 70,643) | Cost-Saving |
| tNGS<br>(3 sites, \$148) | \$1.6m<br>(\$ 1,607,618-\$ 1,608,106) | | \$671 |
| tNGS<br>(4 sites, \$171) | \$1.8m<br>(\$1,857,699 - \$1,858,282) | | \$1,283 |
| tNGS<br>(6 sites, \$215) | \$2.3m<br>(\$2,335,511 - \$2,336,233) | | \$2,453 |

Figure 2 presents the results of the sensitivity analyses. Panel A shows that higher tNGS sensitivity to bedaquiline results in fewer DALYs overall. Regardless of DST algorithm, as bedaquiline resistance increases, the total DALYs also increase. At the lower-bound of considered tNGS sensitivity values for bedaquiline (68%) ^15^, bedaquiline resistance would need to be ≥11% for tNGS to outperform the SOC. At higher sensitivity values, the tNGS algorithm results in fewer DALYs than the SOC, regardless of bedaquiline prevalence. Panel B illustrates the impact of increased access to DST results through tNGS on total DALYs averted at different levels of tNGS decentralisation. As access to DST improves, the number of DALYs averted through tNGS increases, reaching up to 1101 DALYs averted for a 20% increase in access. Subsequently, per level of decentralisation, as access increases the incremental cost per DALY averted decreases: for example, at a 20% improvement in access the incremental cost per DALY averted now ranges from $433-$1094 (for 3 and 6 sites respectively).

**Figure 2:**
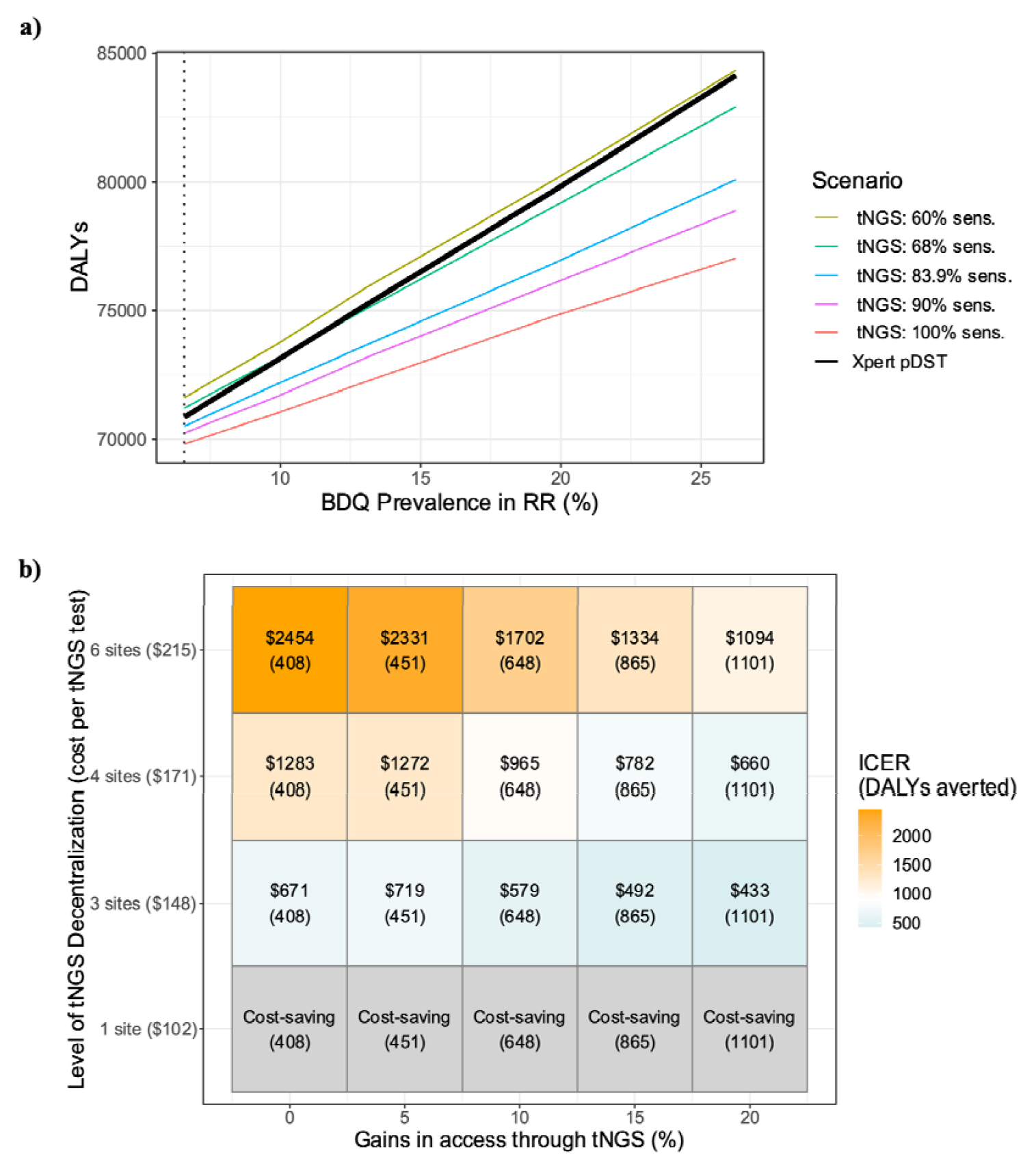
Results of sensitivity analyses. Panel A: Impact of prevalence of BDQ resistance and tNGS sensitivity to BDQ on total DALYs. Panel B: Impact of increased DST coverage through tNGS on incremental cost per DALY averted at different levels of tNGS decentralisation

## DISCUSSION

The modelling analysis demonstrates that implementing tNGS for DST could improve outcomes for RR/MDR-TB in South Africa. Under the assumed context (∼6.5% BDQ resistance, 0% difference in DST access between algorithms, and a tNGS TAT of 10.5 days), tNGS resulted in a higher proportion of resistance profiles being correctly identified and returned as compared to SOC. tNGS also yielded a modest increase in survival rate (+0.2%) and averted 408 DALYs. Furthermore, tNGS returned results faster than the SOC, enabling quicker regimen adjustments for patients and reducing time spent on ineffective treatment.

This reduction in delays contributed to lower mortality rates and better health outcomes for the patient and could potentially reduce the amplification of resistance to other drugs in the regimen. Additionally, DST through tNGS shortened the overall infectious period (from diagnosis to effective treatment initiation), thereby potentially decreasing transmission potential among individuals with RR/MDR TB during the DST period. Consequently, tNGS not only improves individual patient outcomes but could offer public health benefits by reducing the spread of drug-resistant TB cases.

From a cost perspective, a fully centralized tNGS DST algorithm, with testing conducted at a single site, is estimated to cost less per test as compared to the SOC (tNGS: $102, SOC: $137). Under this centralized scenario, tNGS is both cost-saving and associated with better patient outcomes, highlighting it as a feasible option for DST in South Africa. Compared to the costs estimated by Shrestha et al ^17^ ($197/test compared to our $102/test), the lower cost in our analysis is likely attributable to higher estimated volumes per batch in a centralized model, larger batch sizes in the equipment modelled and overall reductions in technology costs.

When considering tNGS decentralisation in South Africa, each additional testing site results in cost increases per individual tested. These cost increases stem from the need for dedicated equipment and personnel at each site. Moreover, as the number of sites increases, the total volume of tNGS tests performed per site decreases. Given that we have modelled tNGS to minimise TAT, these reduced testing volumes result in higher costs as tNGS runs are not being performed at full capacity. This results in higher costs per DALY averted, ranging from $671-$2,454 for 3-6 sites respectively, which could be considered cost-effective under some commonly used willingness to pay thresholds ($900-$3600) ^28–30^.

The sensitivity analyses highlight the potential impact of increased DST access and improved bedaquiline resistance sensitivity on DALYs. For the base case, the model assumes no difference in access to DST between algorithms. If, in practice, tNGS improves the return and use of accurate DST results, the total DALYs averted would increase, and the cost per DALY averted would decrease. This is particularly relevant as decentralisation levels increase. For example, a 20% increase in access could reduce the cost per DALY averted by 53%, emphasizing that implementation approaches that increase access may improve the feasibility of decentralization. Furthermore, it is likely that there will be improvements in tNGS sensitivity to bedaquiline resistance over time as updates to the catalogue of mutations take place^31^. This could further reduce DALYs and lower the cost per DALY averted, enhancing the case for decentralised tNGS implementation. Lastly, although not modelled in this work, a potential cost-saving mechanism for tNGS would be cost-sharing or capacity sharing of tNGS equipment across multiple healthcare programmes. Conducting tNGS in this manner would likely require notable co-ordination between programmes but would ensure better batch utilization and hence would reduce overall per test costs.

There are several limitations to this modelling work. First, assumptions about tNGS implementation and TAT have been made. While these have been informed by in-country experts, there is potential that tNGS may have a different TAT than what is modelled or that there may be difficulties in introducing this technology in a routine manner. If routine implementation challenges result in delayed TAT beyond that modelled in this work, it could affect both its cost and impact compared to SOC. Once implementation begins, it will be critical to revisit the modelling process using observed data and lessons learned to generate updated estimates of cost and impact. Similarly, in this work assumptions around reduced delays leading to lower mortality rates and better outcomes have been made. Further empiric evidence would be needed to confirm that these hold true in the context of DST through tNGS in South Africa. Additionally, this analysis modelled only a subset of key resistance types routinely considered for RR/MDR TB ^12^. It does not account for potential gains of tNGS for detecting other resistance types, such as isoniazid, which may be missed with the initial diagnostic. While the impact cannot yet be quantified, it is important to note that the introduction of tNGS systems would be a sustainable, or “future-proof”, methodology for DST. Unlike current methods, which require the development of new tools for each additional drug, tNGS systems can easily be updated to include new or emerging resistance targets, providing long-term adaptability, cost-savings, and a decrease in time between when a new diagnostic target is needed and when it is available.

Lastly, this analysis is centred on the South African context. Prevalence rates for drug resistance, the modelled DST algorithms, treatment initiation processes, instrument choice and cost estimates may not be generalizable to other countries. Countries with different drug resistance profiles, infrastructure (both for testing and return of results), or algorithms for DST may reach different conclusions regarding feasibility and cost-effectiveness. Further we only included Illumina instruments in our analysis as these were the instruments that NICD were considering in their programme, however, future analyses should include other manufacturers and products that have competitive costs, especially for low volumes. Importantly, South Africa has a robust specimen transport network, enabling a fully centralized DST system with negligible per sample costs-which may be infeasible or challenging in other countries without significant infrastructure investment^32^. Conducting similar analyses in other countries or across various country archetypes would provide a more comprehensive understanding of the global applicability of tNGS for DST.

## CONCLUSIONS

This analysis highlights centralised tNGS as a promising alternative method for DST in South Africa. tNGS demonstrated the potential to reduce DALYs, shorten time to results, and decrease overall infectious time while yielding a higher number of correct results compared to the SOC. Centralised tNGS has the potential to be cost-saving and while decentralisation increases costs notably, decentralised may still be considered cost-effective. Future work should focus on refining assumptions about tNGS implementation based on real-world observations from in-country rollouts. Expanding the analysis to include DST in other countries or across diverse healthcare setting would provide valuable insights into the broader feasibility and cost-effectiveness of tNGS globally.

## Data Availability

All data produced in the present study are available upon reasonable request to the authors

## AUTHOR CONTRIBUTIONS

AdN, BEN, KG, SO, NI, AS, SK and SG conceived and designed the analysis. NI and SO supplied and collected the data. AdN, SG and TO conducted the analysis. BEN, NI, SO and SG reviewed and verified the data. Analysis interpretation was completed by AdN, TO, BEN, SG, NI, SO, AS and KG and results and analysis were validated by AZ, SS, TR, RC and SU. The paper was written by AdN, TO, BEN and SG and reviewed and edited by AZ, SK,SS, AS, SJ, NI, SO, RC, SU, TR and KG. All authors were permitted access to all data in this analysis and accept responsibility for submitting the manuscript for publication.

## ETHICS COMMITTEE APPROVAL

No ethics approval was required for this work.

## DATA SHARING

Model results can be shared upon request.

## REFERENCES

1 World Health Organization. Global tuberculosis report 2024. Geneva, 2024 https://iris.who.int/bitstream/handle/10665/379339/9789240101531-eng.pdf?sequence=1 (accessed Nov 17, 2024).

2 South African National Department of Health, Centre for Tuberculosis incorporating the National TB Reference Laboratory National Institute for Communicable Diseases. Webinar: BPaL-L and the emergence of bedaquiline resistance. 2023; published online March. https://knowledgehub.health.gov.za/system/files/2024-03/In-Session%20slides_BDQ%20resistance%20webinar.pdf (accessed Nov 17, 2024).

3 World Health Organization. Tuberculosis Profile: South Africa. 2023. https://worldhealthorg.shinyapps.io/tb_profiles/?_inputs_&tab=%22charts%22&lan=%22EN%22&iso2=%22ZA%22&entity_type=%22country%22 (accessed Nov 18, 2024).

4 Centre for Tuberculosis National Institute of Communicable Diseases a division of the National Health Laboratory Service. NICD Data and Communication. 2024; published online Oct 1.

5 Ismail NA, Mvusi L, Nanoo A, et al. Prevalence of drug-resistant tuberculosis and imputed burden in South Africa: a national and sub-national cross-sectional survey. Lancet Infect Dis 2018; 18: 779.

6 Nicholson TJ, Hoddinott G, Seddon JA, et al. A systematic review of risk factors for mortality among tuberculosis patients in South Africa. Syst Rev 2023; 12: 23.

7 World Health Organization. WHO consolidated guidelines on tuberculosis Module 4: Treatment Drug-resistant tuberculosis treatment 2022 update. Geneva, 2022 https://iris.who.int/bitstream/handle/10665/365308/9789240063129-eng.pdf?sequence=1 (accessed Jan 20, 2025).

8 Dean AS, Tosas Auguet O, Glaziou P, et al. 25 years of surveillance of drug-resistant tuberculosis: achievements, challenges, and way forward. Lancet Infect Dis 2022; 22: e191–6.

9 Dheda K, Gumbo T, Maartens G, et al. The epidemiology, pathogenesis, transmission, diagnosis, and management of multidrug-resistant, extensively drug-resistant, and incurable tuberculosis. Lancet Respir Med 2017; 5: 291–360.

10 Cepheid. Xpert® MTB/RIF Ultra. https://www.cepheid.com/en-GB/tests/tb-emerging-infectious-diseases/xpert-mtb-rif-ultra.html (accessed Sept 27, 2023).

11 Cepheid. Xpert® MTB/XDR. 2024. https://www.cepheid.com/en-NL/tests/tb-emerging-infectious-diseases/xpert-mtb-xdr.html (accessed Nov 28, 2024).

12 South African National Department of Health. Clinical Management of Rifampicin Resistant Tuberculosis: Updated Clinical Reference Guide. 2023 https://www.health.gov.za/wp-content/uploads/2023/10/Updated-RR-TB-Clinical-Guidelines-September-2023.pdf (accessed Nov 28, 2024).

13 Liebenberg D, Gordhan BG, Kana BD. Drug resistant tuberculosis: Implications for transmission, diagnosis, and disease management. Front Cell Infect Microbiol 2022; 12: 943545.

14 World Health Organization. Use of targeted next-generation sequencing to detect drug-resistant tuberculosis: rapid communication, July 2023. 2023 https://iris.who.int/bitstream/handle/10665/371687/9789240076372-eng.pdf?sequence=1 (accessed Nov 28, 2024).

15 World Health Organization. WHO Operational handbook on tuberculosis. Module 3: diagnosis - rapid diagnostics for tuberculosis detection, third edition. Geneva, 2024 https://iris.who.int/bitstream/handle/10665/376155/9789240089501-eng.pdf?sequence=1 (accessed Nov 28, 2024).

16 Iyer A, Ndlovu Z, Sharma J, et al. Operationalising targeted next-generation sequencing for routine diagnosis of drug-resistant TB. Public Health Action 2023; 13: 43.

17 Shrestha S, Addae A, Miller C, Ismail N, Zwerling A. Cost-effectiveness of targeted next-generation sequencing (tNGS) for detection of tuberculosis drug resistance in India, South Africa and Georgia: a modeling analysis. EClinicalMedicine 2025; 79: 103003.

18 Colman RE, Seifert M, De la Rossa A, et al. Evaluating culture-free targeted next-generation sequencing for diagnosing drug-resistant tuberculosis: a multicentre clinical study of two end-to-end commercial workflows. Lancet Infect Dis 2024; 0. DOI:10.1016/S1473-3099(24)00586-3/ATTACHMENT/596E7DA2-B966-45BB-B847-EA3E01E7EA0B/MMC1.PDF.

19 World Health Organization. WHO consolidated guidelines on tuberculosis. Module 3: diagnosis-rapid diagnostics for tuberculosis detection,2021 update. World Health Organization 2021; Module 3: 1–164.

20 TreeAge LLC. TreeAgePro Healthcare Software. 2023. https://www.treeage.com/ (accessed Nov 17, 2024).

21 Posit Software. Download RStudio | The Popular Open-Source IDE from Posit. 2024. https://posit.co/products/open-source/rstudio/ (accessed Dec 18, 2024).

22 National health laboratory Service (NHLS). NHLS Annual Report 2021-2022. 2022 https://www.nhls.ac.za/wp-content/uploads/2022/10/NHLS_AR_2022_web_version.pdf (accessed March 4, 2024).

23 South African National Department of Health. CLINICAL MANAGEMENT OF RIFAMPICIN-RESISTANT TUBERCULOSIS: Updated Clinical Reference Guide. 2023.

24 Reidpath DD, Allotey PA, Kouame A, Cummins RA. Measuring health in a vacuum: examining the disability weight of the DALY. Health Policy Plan 2003; 18: 351–6.

25 Illumina Inc. NextSeq 1000 & 2000 System Specifications. https://emea.illumina.com/systems/sequencing-platforms/nextseq-1000-2000/specifications.html (accessed Jan 21, 2025).

26 GenoScreen. Deeplex ® Myc-TB: User Manual. 2022.

27 Cassim N, Omar SV, Masuku SD, et al. Xpert MTB/XDR implementation in South Africa: cost outcomes of centralised vs. decentralised approaches. IJTLD OPEN 2024; 1: 215.

28 Meyer-Rath G, Van Rensburg C, Larson B, Jamieson L, Rosen S. Revealed willingness-to-pay versus standard cost-effectiveness thresholds: Evidence from the South African HIV Investment Case. PLoS One 2017; 12: e0186496.

29 Edoka IP, Stacey NK. Estimating a cost-effectiveness threshold for health care decision-making in South Africa. Health Policy Plan 2020; 35: 546–55.

30 Ochalek J, Lomas J, Claxton K. Estimating health opportunity costs in low-income and middle-income countries: a novel approach and evidence from cross-country data. BMJ Glob Health 2018; 3: 964.

31 Walker TM, Fowler PW, Knaggs J, et al. The 2021 WHO catalogue of Mycobacterium tuberculosis complex mutations associated with drug resistance: a genotypic analysis. Lancet Microbe 2022; 3: e265.

32 Girdwood SJ, Crompton T, Cassim N, et al. Optimising courier specimen collection time improves patient access to HIV viral load testing in South Africa. Afr J Lab Med 2022; 11: 1725.

33 Ahmad D, Morgan WKC, Schwartzman K, Menzies D. How long are TB patients infectious? CMAJ: Canadian Medical Association Journal 2000; 163: 157.

34 World Health Organization Regional Office fro South-East Asia. Frequently Asked Questions about tuberculosis. New Delhi, 2013 https://iris.who.int/bitstream/handle/10665/205081/B5009.pdf?sequence=1%26isAllowed=y (accessed Dec 13, 2024).

35 Menzies NA, Quaife M, Allwood BW, et al. Lifetime burden of disease due to incident tuberculosis: a global reappraisal including post-tuberculosis sequelae. Lancet Glob Health 2021; 9: e1679–87.

36 Salomon JA, Vos T, Hogan DR, et al. Common values in assessing health outcomes from disease and injury: Disability weights measurement study for the Global Burden of Disease Study 2010. The Lancet 2012; 380: 2129–43.

37 James SL, Abate D, Abate KH, et al. Global, regional, and national incidence, prevalence, and years lived with disability for 354 Diseases and Injuries for 195 countries and territories, 1990-2017: A systematic analysis for the Global Burden of Disease Study 2017. The Lancet 2018; 392: 1789–858.

38 Institute for Health Metrics and Evaluation (IHME). Global Burden of Disease 2021: Findings from the GBD 2021 Study. Seattle,WA, 2024 https://www.healthdata.org/research-analysis/library/global-burden-disease-2021-findings-gbd-2021-study (accessed Sept 13, 2024).

39 Stats SA. Mid-Year population Estimates. 2024 https://www.statssa.gov.za/?page_id=1854&PPN=P0302&SCH=73952 (accessed Nov 5, 2024).

40 World Health Organization. Global tuberculosis report 2023. Geneva, 2023 https://iris.who.int/bitstream/handle/10665/373828/9789240083851-eng.pdf?sequence=1 (accessed Feb 16, 2024).

41 Ndjeka N, Campbell JR, Meintjes G, et al. Treatment outcomes 24 months after initiating short, all-oral bedaquiline-containing or injectable-containing rifampicin-resistant tuberculosis treatment regimens in South Africa: a retrospective cohort study. Lancet Infect Dis 2022; 22: 1042.

42 World Health Organization. Genomics costing tool: user manual. Geneva, 2024 https://www.who.int/publications/i/item/9789240090866 (xaccessed Jan 24, 2025).

43 Illumina. Illumina Global Health Access Pricing. 2024. https://sapac.illumina.com/destination/global-health-access-initiative.html?media=9080385&utm_medium=Other_Campaigns&catt=Other_Campaigns_Press_Release (accessed Jan 24, 2025).

