## Supplemental Materials for "Cost-effectiveness of Targeted Next Generation Sequencing for TB drug-resistance testing as an alternative to the standard of care in South Africa"

This appendix provides relevant parameter values and sources for the model and costing as well as further detail on certain calculations and assumptions.

**Model Parameters and Sources**

Table S1 provides the relevant model parameters, description, values and sources used to parametrise the decision tree model for both the standard of care (SOC) and targeted next generation sequencing (tNGS) drug susceptibility testing (DST) algorithms.

| **Table S1: Model parameter values** | | | |
| --- | --- | --- | --- |
| **Name** | **Description** | **Value** | **Source** |
| **Costs** | | | |
| cpDST | Cost per sample test in pDST | $97 | Calculated with data and guidance from NICD (see Table S5, S6)  ^1^ |
| ctNGS (1 Site) | Cost per sample test in tNGS | $102 |  |
| ctNGS (3 Sites) | Cost per sample test in tNGS | $202 |  |
| ctNGS (4 Sites) | Cost per sample test in tNGS | $267 |  |
| ctNGS (6 Sites) | Cost of per sample test in tNGS | $541 |  |
| cXpertXDR | Cost Xpert XDR test | $40 |  |
| **Prevalence Drug Resistance** | | | |
| pBDQFQRR | Prevalence of BDQ resistant among FQ and R resistant | 0.17 | ^2^ |
| pBDQRR | Prevalence of BDQ resistant among RR | 0.05 |  |
| pFQRR | Prevalence of FQ resistant among RR | 0.13 |  |
| pLZDBDQFQRR | Prevalence of LZD resistant among MDR with BDQ resistant | 0.065 |  |
| pLZDBDQRR | Prevalence of LZD resistant with BDQ and R resistant | 0.045 |  |
| pLZDFQRR | Prevalence of LZD resistant among FQ and R resistant | 0.026 |  |
| pLZDRR | Prevalence of LZD resistant among RR | 0.026 |  |
| pRif | Prevalence of Rif resistance in TB + population | 0.051 | ^1^ |
| pIMR | Prevalence ING mono-resistance | 0.067 | ^3^ |
| **Test Performance** | | | |
| SenBdqpDST | Sensitivity of pDST to detect BDQ resistant | 0.998316498 | ^2^ |
| SenFQpDST | Sensitivity of pDST to detect FQ resistant | 0.991997534 |  |
| SenLZDpDST | Sensitivity of pDST to detect LZD resistant | 0.99537037 |  |
| SenFQXpertXDR | Sensitivity of Xpert XDR to detect FQ resistant | 0.93 |  |
| SenRifXpert | Sensitivity of Xpert to detect rif resistant | 0.949 |  |
| SpeBdQpDST | Specificity of pDST on BDQ Resistant | 0.997996633 |  |
| SpeFQpDST | Specificity of pDST on FQ resistant | 0.999001233 |  |
| SpeLZDpDST | Specificity of pDST on LZD resistant | 1 |  |
| SpeFQXpertXDR | Specificity of Xpert XDR to detect FQ resistant | 0.98 |  |
| SpeRifXpert | Specificity of Xpert to R/o Rif resistant | 0.991 |  |
| SenBdqtNGS | Sensitivity of tNGS to detect Bdq resistant | 0.839 | ^4^ |
| SenFQtNGS | Sensitivity of tNGS to detect FQ resistant | 0.966 |  |
| SenLZDtNGS | Sensitivity of tNGS to detect LZD resistant | 0.462 |  |
| SenInhtNGS | Sensitivity of tNGS to diagnose INH resistant | 0.96 |  |
| SenRiftNGS | Sensitivity of tNGS to detect Rif resistant | 0.973 |  |
| SpeBdQtNGS | Specificity of tNGS for BDQ | 0.977 |  |
| SpeFQtNGS | Specificity of tNGS for FQ | 0.985 |  |
| SpeLZDtNGS | Specificity of tNGS on LZD | 0.998 |  |
| SpeInhtNGS | Specificity of tNGS to rule out INH resistant | 0.97 |  |
| SpeRiftNGS | Specificity of tNGS to diagnose Rif resistant | 1 |  |
| **Contamination and repeat Rates** | | | |
| pContDST | Probability of contamination and LTFU in pDST | 0.14 | ^1^ |
| pIndtNGS | Probability of Indeterminate results from tNGS | 0.15 |  |
| pRepeatpDST | Prob. of repeat test in pDST | 0.5 | Assumption |
| pRepeattNGS | probability of repeat test tNGS | 0.8 | ^1^ |
| pIndXpertXDR | probability of indeterminant result Xpert XDR | 0.0296 | ^2^ |
| pRepeatXpertXDR | Probability of repeat test in Xpert XDR after indeterminant result | 0.1 |  |
| **Treatment initiation and completion** | | | |
| pRXinitiation | Probability of treatment initiation | 0.9000157 | ^2^ |
| pINDRXcompletion | Probability of Individualized treatment completion | 0.73 |  |
| pHRZEcomp | Probability of completion of HRZE | 0.9 |  |
| pBPaLcompletion | Probability of completion of BPaL | 0.9 |  |
| **Mortality** | | | |
| pDeathBPAL | Probability of death in BPAL regimen | 0.06741573 | ^2^ |
| pDeathBPAL-L | Probability of death in BPAL-L regimen | 0.06741573 |  |
| pDeathincRX | Probability of death among incomplete and wrong treatment | 0.485527411 |  |
| pDeathindRx | Probability of death in Individualized Regimen | 0.125 |  |
| pDeathnoRx | Probability of death with no treatment | 1 |  |
| pDeathHRZERX | Probability of death in HRZE regimen | 0.04 |  |
| pDeathHR | Probability of death in HR regimen | 0.04 |  |
| **Timing** | | | |
| time_tngs | TAT tNGS | 10.5 days | ^1^ |
| time_pdst | TAT pDST | 56 days |  |
| time_dst_inc | Assumed time penalty for missing or incorrect DST | 84 days | Assumption |
| time_culture_check | TAT of month 3 culture results (3 month sample request, 2 month TAT) | 140 days | ^5,6^ |
| time_treatment_effect | Time taken until individual on correct treatment no longer infectious | 14 days | ^7,8^ |
| time_no_treatment | Expected survival time if no treatment taken | 2.5 years | ^9^ |
| time_profile | Either time_tngs or time_dst depending on algorithm | | |
| time_regimen_change | 28 days (DST – next monthly visit), 3.5 days (tNGS, 2-week visit) | | |
| **Disability Weights and DALYs** | | | |
| dw_treated | Disability weight for treated TB while on effective treatment | 0.051 | ^10,11^ |
| dw_untreated | Average disability weight for untreated TB | 0.384 | ^12^ |
| daly_deaths | DALYs associated with a death in the model  (Calculated using median age of TB and SA life expectancy) | 31.5 | ^13,14^ |

**Mortality Assumptions**

The mortality assumptions used in this analysis follow closely those presented in the work by Shrestha et al ^2^ and are presented in Table S2. An additional assumption which has been included is that of modelling additional early deaths as a result of delayed switching to individualised regimens. Here this is assumed to only apply to those individuals who require a long individualised regimen and would be correctly switched based on the resistance results returned. This mortality is assumed to be at a consistent rate per month on ineffective treatment. This mortality is taken to be approximately 5.18% (41% of the assumed mortality on individualised regimens) - based on estimations from Ndjeka et al that estimate for the BPAL regimen that approximately 41% of TB deaths (at 18 months) were recorded as “early” (within 3 months of starting treatment) ^15^.

| **Table S2: Model mortality assumptions** | | | | | |
| --- | --- | --- | --- | --- | --- |
| **Treatment Status** | **Correct Regimen** | **Resulting Regimen** | **Resistance Profile Correct** | **Mortality parameter*** | **Modelling for potential death before regimen switch **** |
| No treatment | - | - | - | pDeathnoRx | No |
| Incomplete Treatment | - | - | - | pDeathincRx | No |
| Complete Treatment | BPaL/  BPaL-L | BPaL/  BPaL-L | - | pDeathBPAL | No |
| Complete Treatment | BPaL/  BPaL-L | Individual | No | pDeathindRx | No |
| Complete Treatment | Individual | Individual | Yes | pDeathindRx | Yes |
| Complete Treatment | Individual | Individual | No | pDeathincRx | No |
| Complete Treatment | Individual | BPaL/  BPaL-L | No | pDeathincRx | No |
| * Parameter used to represent the likelihood of death at 18 months after receiving diagnosis (if no or incomplete treatment) or starting an effective treatment  ** Represents where additional mortality is incorporated in cases of delayed turnaround of results when a regimen switch is required and occurs correctly | | | | | |

**Calculation of Infectious Time**

Infectious time post diagnosis is estimated as the sum of the time on ineffective treatment and the time in which it takes appropriate treatment to have an effect (assumed 14 days^7,8^). This is highlighted by treatment status, correct and determined regimen and resistance results in Table S3 below. Here time spent on ineffective treatment is based, where relevant, on the time take to return DST results and the subsequent time required to switch regimens (likely at the next scheduled visit). Individuals started on a BPaL regimen who require a BPaL regimen as such would have no time on ineffective treatment. Lastly, individuals who should receive a long-individualised regimen but do not receive resistance results indicating this are assumed to be switched at a later date, after the 3-month culture indicates failing treatment.

| **Table S3: Infectious time calculations** | | | | |
| --- | --- | --- | --- | --- |
| **Treatment Status** | **Correct Regimen** | **Resulting Regimen** | **Resistance Profile Correct** | **Infectious Time Calculation** |
| No treatment | - | - | - | Time_no_treatment |
| Incomplete Treatment | - | - | - | Time_no_treatment |
| Complete Treatment | BPaL/  BPaL-L | BPaL/  BPaL-L | - | Time_treatment_effect |
| Complete Treatment | BPaL/  BPaL-L | Individual | No | Time_treatment_effect |
| Complete Treatment | Individual | Individual | Yes | Time_treatment_effect + time_profile + time_regimen_change |
| Complete Treatment | Individual | Individual | No | Time_culture_check + time_treatment_effect |
| Complete Treatment | Individual | BPaL/  BPaL-L | No | Time_culture_check + time_treatment_effect |

**DALY Calculations**

The equations used to calculate the DALYs per individual, based on their treatment status, survival status, true resistance profile, resulting resistance profile and DST algorithm utilised are provided in Table S4 below. DALYs were considered in terms of years of life-lost (YLL) for each modelled death and in terms of years lived with disability (YLD) ^16^. Given that the model is not age-structured, YLL per death were estimated as the difference between the estimated life-expectancy for South Africa in 2024 (66.5 years) ^13^ and the median age of TB (estimated at 35 years in based on WHO TB incidence data for South Africa^17^). This resulted in a calculated YLL of 31.5 years per death. YLDs were estimated on a group-by-group case where each group represents a unique combination of true resistance profile, determined resistance profile and treatment regimen, treatment initiation status, treatment completion status and survival status as well as timing elements relevant to each DST algorithm. In each case, the YLD calculation uses treated and untreated disability weights (DW) applied to the relevant time in each state, with time per states based on the make-up of the group ^16^.

| **Table S4: DALY Calculations per individual** | | | | | |
| --- | --- | --- | --- | --- | --- |
| **Treatment Status** | **Correct Regimen** | **Resulting Regimen** | **Resistance Profile Correct** | **Survival** | **DALY (per person)** |
| No/incomplete treatment | - | - | - | Yes | Dw_untreated*time_no_treatment |
| No/incomplete treatment | - | - | - | No | Daly_deaths+Dw_untreated*time_no_treatment |
| Complete Treatment | BPaL/  BPaL-L | BPaL/  BPaL-L | - | Yes | Dw_untreated*time_treatment_effect + dw_treated*(time_bpal-time_treatment_effect) |
| Complete Treatment | BPaL/  BPaL-L | BPaL/  BPaL-L | - | No | Dw_untreated*time_treatment_effect + dw_treated*(time_bpal-time_treatment_effect)+daly_deaths |
| Complete Treatment | BPaL/  BPaL-L | Individual | No | Yes | Dw_untreated*(time_treatment_effect+profile_time+time_reg_change) + Dw_treated*(time_individual-time_treatment_effect) |
| Complete Treatment | BPaL/  BPaL-L | Individual | No | No | Dw_untreated*(time_treatment_effect+profile_time+time_reg_change) + Dw_treated*(time_individual-time_treatment_effect))+daly_deaths |
| Complete Treatment | Individual | Individual | Yes | Yes | Dw_untreated*(time_treatment_effect+profile_time+time_reg_change) + dw_treated*(*(time_individual-time_treatment_effect) |
| Complete Treatment | Individual | Individual | Yes | No | Dw_untreated*(time_treatment_effect+profile_time+time_reg_change) + dw_treated*(*(time_individual-time_treatment_effect)+daly_deaths |
| Complete Treatment | Individual | BPaL/  BPaL-L | No | Yes | Dw_untreated*(time_culture_check + time_treatment) + dw_treated*(time_individual-time_treatment_effect) |
| Complete Treatment | Individual | BPaL/  BPaL-L | No | No | Dw_untreated*(time_culture_check + time_treatment) + dw_treated*(time_individual-time_treatment_effect)+ daly_deaths |
| Complete Treatment | Individual | Individual | No | Yes | Dw_untreated*(time_culture_check + time_treatment) + dw_treated*(time_individual-time_treatment_effect) |
| Complete Treatment | Individual | Individual | No | No | Dw_untreated*(time_culture_check + time_treatment) + dw_treated*(time_individual-time_treatment_effect)+ daly_deaths |

**Costing of tNGS and SOC algorithms**

**Table S5: Cost per test stratified by cost category for the standard of care and tNGS testing algorithms for 10,000 tests**

| **Standard-of-care** | | |
| --- | --- | --- |
| **Cost category** | **Cost per test** | **Notes/sources** |
| **Culture and pDST** | | |
| **Consumables** | **$23** | Consumable costs for culture and pDST are provided by the NICD. ^1^ |
| **Equipment** | **$65** | Equipment costs are provided entirely by the NICD and are annualized using a 3% discount rate and a 10-year working lifespan. The number of items required per site was determined and multiplied by the 15 DST/culture sites in operation nationally. Prices were inclusive of importation duties/taxes where relevant.^1^ |
| **Staff** | **$8** | Staff and their salaries for all activities related to culture and pDST are sourced from the NICD. The hands-on staff time to conducted different activities associated with culture and pDST was determined per batch to determine a staff cost per test. ^1^ |
| **Energy** | **$0.3** | Energy usage per batch per instrument and cost per kWh was used to determine the cost of running GeneXpert and MGIT960 instruments. ^1^ |
| **Xpert XDR** | **$39.90** | The all-inclusive Xpert XDR cost from Cassim et al, inflated to 2024, is used^18^. |
| **Total SOC** | **$137** | |
| **tNGS (1 site)** | | |
| **Consumables** | **$78** | Consumables include all items required for DNA preparation and PCR and are sourced from the NICD. ^1^ |
| **Cartridge** | **$6** | The sequencing cartridge cost is sourced from Illumina's Global Health Access Pricing, while cartridge prices vary based on instrument utilization, with significant fluctuations depending on the number of sites. |
| **Equipment** | **$14** | Site-specific equipment is sourced from the WHO Genomics Tool ^19^ and discussions with NICD partners^1^. Sequencing platforms (e.g., NextSeq 1000) are procured through Illumina's Global Health Access Pricing^20^. All items are annualized using a 3% discount rate, with the working lifespan varying depending on the equipment. |
| **Staff** | **$3** | Staff types are sourced from the NICD, and the time spent on each PCR workflow step is obtained from the Genoscreen manual^21^. |
| **Bioinformatics/ energy/quality assurance** | **$1** | Bioinformatics, energy and quality assurance are all provided by the NICD. |
| **Total tNGS** | **$102** | |

**Table S6: Cost per test for tNGS for different levels of decentralization**

| **Variable** | Scenario 1 | Scenario 2 | Scenario 3 | Scenario 4 |
| --- | --- | --- | --- | --- |
| Annual tNGS test volume | 10,000 | 10,000 | 10,000 | 10,000 |
| Number of sites | 1 | 3 | 4 | 6 |
| Objective | Minimise cost | Minimise TAT | Minimise TAT | Minimise TAT |
| Sequencing TAT (days) | 7 | 7 | 7 | 7 |
| Sequencing cartridge cost per test | $6 | $24 | $32 | $48 |
| Total annual cost | $1,017,883 | $1,484,326 | $1,707,643 | $2,154,277 |
| Cost per test | $102 | $148 | $171 | $215 |

Site-specific volume distributions

The level of decentralization significantly influences the cost per test for tNGS. As shown in Table S7, test volumes are not distributed equally across sites due to the location of the sites and the way the sample transportation is organized. This variability can lead to higher costs due to low instrument utilization. For instance, with full decentralization (six testing sites), the site with the lowest volume handles only 6% of the total testing volume, while the highest-volume site accounts for 25%. Since instruments are operated weekly in order to keep within a 7-day sequencing TAT, the instrument at the lowest-volume site processes very few samples, resulting in higher costs per test at that site and a higher average cost per test at all sites.

**Table S7: Site specific test distributions for each level of decentralization**

| 1 site | 2 sites | 3 sites | 4 sites | 5 sites | 6 sites |
| --- | --- | --- | --- | --- | --- |
| 100% | 53% | 29% | 29% | 20% | 14% |
|  | 47% | 47% | 25% | 25% | 25% |
|  |  | 23% | 23% | 23% | 23% |
|  |  |  | 23% | 23% | 23% |
|  |  |  |  | 9% | 9% |
|  |  |  |  |  | 6% |

**REFERENCES**

1	Centre for Tuberculosis National Institute of Communicable Diseases a division of the National Health Laboratory Service. NICD Data and Communication. 2024; published online Oct 1.

2	Shrestha S, Addae A, Miller C, Ismail N, Zwerling A. Cost-effectiveness of targeted next-generation sequencing (tNGS) for detection of tuberculosis drug resistance in India, South Africa and Georgia: a modeling analysis. *EClinicalMedicine* 2025; 79: 103003.

3	Ismail NA, Mvusi L, Nanoo A, *et al.* Prevalence of drug-resistant tuberculosis and imputed burden in South Africa: a national and sub-national cross-sectional survey. *Lancet Infect Dis* 2018; 18: 779.

4	Colman RE, Seifert M, De la Rossa A, *et al.* Evaluating culture-free targeted next-generation sequencing for diagnosing drug-resistant tuberculosis: a multicentre clinical study of two end-to-end commercial workflows. *Lancet Infect Dis* 2024; 0. DOI:10.1016/S1473-3099(24)00586-3/ATTACHMENT/596E7DA2-B966-45BB-B847-EA3E01E7EA0B/MMC1.PDF.

5	South African National Department of Health, Centre for Tuberculosis incorporating the National TB Reference Laboratory National Institute for Communicable Diseases. Webinar: BPaL-L and the emergence of bedaquiline resistance. 2023; published online March. https://knowledgehub.health.gov.za/system/files/2024-03/In-Session%20slides_BDQ%20resistance%20webinar.pdf (accessed Nov 17, 2024).

6	South African National Department of Health. Clinical Management of Rifampicin Resistant Tuberculosis: Updated Clinical Reference Guide. 2023 https://www.health.gov.za/wp-content/uploads/2023/10/Updated-RR-TB-Clinical-Guidelines-September-2023.pdf (accessed Nov 28, 2024).

7	Ahmad D, Morgan WKC, Schwartzman K, Menzies D. How long are TB patients infectious? *CMAJ: Canadian Medical Association Journal* 2000; 163: 157.

8	World Health Organization Regional Office fro South-East Asia. Frequently Asked Questions about tuberculosis. New Delhi, 2013 https://iris.who.int/bitstream/handle/10665/205081/B5009.pdf?sequence=1%26isAllowed=y (accessed Dec 13, 2024).

9	Menzies NA, Quaife M, Allwood BW, *et al.* Lifetime burden of disease due to incident tuberculosis: a global reappraisal including post-tuberculosis sequelae. *Lancet Glob Health* 2021; 9: e1679–87.

10	Salomon JA, Vos T, Hogan DR, *et al.* Common values in assessing health outcomes from disease and injury: Disability weights measurement study for the Global Burden of Disease Study 2010. *The Lancet* 2012; 380: 2129–43.

11	James SL, Abate D, Abate KH, *et al.* Global, regional, and national incidence, prevalence, and years lived with disability for 354 Diseases and Injuries for 195 countries and territories, 1990-2017: A systematic analysis for the Global Burden of Disease Study 2017. *The Lancet* 2018; 392: 1789–858.

12	Institute for Health Metrics and Evaluation (IHME). Global Burden of Disease 2021: Findings from the GBD 2021 Study. Seattle,WA, 2024 https://www.healthdata.org/research-analysis/library/global-burden-disease-2021-findings-gbd-2021-study (accessed Sept 13, 2024).

13	Stats SA. Mid-Year population Estimates. 2024 https://www.statssa.gov.za/?page_id=1854&PPN=P0302&SCH=73952 (accessed Nov 5, 2024).

14	World Health Organization. Global tuberculosis report 2023. Geneva, 2023 https://iris.who.int/bitstream/handle/10665/373828/9789240083851-eng.pdf?sequence=1 (accessed Feb 16, 2024).

15	Ndjeka N, Campbell JR, Meintjes G, *et al.* Treatment outcomes 24 months after initiating short, all-oral bedaquiline-containing or injectable-containing rifampicin-resistant tuberculosis treatment regimens in South Africa: a retrospective cohort study. *Lancet Infect Dis* 2022; 22: 1042.

16	Reidpath DD, Allotey PA, Kouame A, Cummins RA. Measuring health in a vacuum: examining the disability weight of the DALY. *Health Policy Plan* 2003; 18: 351–6.

17	World Health Organization. Tuberculosis Profile: South Africa. 2023. https://worldhealthorg.shinyapps.io/tb_profiles/?_inputs_&tab=%22charts%22&lan=%22EN%22&iso2=%22ZA%22&entity_type=%22country%22 (accessed Nov 18, 2024).

18	Cassim N, Omar SV, Masuku SD, *et al.* Xpert MTB/XDR implementation in South Africa: cost outcomes of centralised vs. decentralised approaches. *IJTLD OPEN* 2024; 1: 215.

19	World Health Organization. Genomics costing tool: user manual. Geneva, 2024 https://www.who.int/publications/i/item/9789240090866 (accessed Jan 24, 2025).

20	Illumina. Illumina Global Health Access Pricing. 2024. https://sapac.illumina.com/destination/global-health-access-initiative.html?media=9080385&utm_medium=Other_Campaigns&catt=Other_Campaigns_Press_Release (accessed Jan 24, 2025).

21	GenoScreen. Deeplex ® Myc-TB: User Manual. 2022.
